# Physical activity and prevalence of constipation in Japanese young people

**DOI:** 10.1101/2024.01.26.24301858

**Authors:** Junichi Watanabe, Shinya Furukawa, Yasunori Yamamoto, Aki Kato, Katsunori Kusumoto, Eiji Takeshita, Yoshio Ikeda, Naofumi Yamamoto, Yuka Saeki, Teruki Miyake, Osamu Yoshida, Yoichi Hiasa

## Abstract

**Background:** Constipation is a very common medical issue among general humans worldwide. The association between physical activity (PA) and constipation is still inconsistent. Additionally, no evidence regarding this issue in young population.

**Aims:** This study aimed to evaluate the association between self-reported physical activity and constipation in Japanese young population, taking the presence or absence of an exercise partner as an additional variable.

**Methods:** The study subjects consisted of 12497 Japanese university students. Information on constipation, exercise frequency, exercise intensity, and exercise partners was obtained from a self-administered questionnaire. Age, sex, body mass index, drinking, smoking, anemia, and sports injury were selected as potential confounding factors.

**Results:** The prevalence of constipation was 6.5% in this cohort. Low, moderate, and high frequency of exercise was independently inversely associated with constipation (adjusted odds ratios [a OR] were low: a OR 0.77 [95% confidence interval (CI) 0.63–0.94], moderate: a OR 0.75 [95% CI 0.60–0.94] and high: a OR 0.70 [95% CI 0.53–0.91], p for trend p=0.002). Low, moderate, and high intensity of exercise was independently inversely associated with constipation (low: a OR 0.77 [95% CI 0.62–0.97], moderate: a OR 0.77 [95% CI 0.63–0.95] and high: a OR 0.70 [95% CI 0.53–0.87], p for trend p=0.001). Exercise with groups and with friends was independently inversely associated with constipation whereas the association between exercising alone and constipation was not significant (groups: a OR 0.70 [95% CI 0.53–0.90] and friends: a OR 0.56 [95% CI 0.42–0.74])

**Conclusion:** In the young Japanese population, frequency and intensity of exercise and presence of exercise partner might be independently inversely associated with constipation.

## Introduction

Constipation is a common gastrointestinal disorder among general humans worldwide. Previous report indicates that the prevalence of constipation ranges from 2.6% to 26.9% [1]. It is estimated that 70–90% of patients with constipation report bothersome symptoms such as bloating, straining, and hard stool persisting for many years, with a negative impact on health-related quality of life [2.3]. Increased physical activity (PA) has long been recommended as part of the management of constipation. The association between PA and constipation has remained inconclusive to date. That is to say, the benefice of increasing daily physical exercise in the treatment of constipation have a lack of evidence. Several studies have showed an inverse association between PA and constipation. In Icelandic, Swedish, Hong Kong, two Chinese and three Japanese studies showed the inverse association between constipation, functional constipation, and PA [4-11]. On the other hand, In US, British, and two Spanish studies were no association between PA and constipation was found [12-15]. Epidemiological evidence on this issue has been limited and inconsistent.

Exercise with others may have a stronger beneficial effect on health compared to exercise alone [16-20]. However, no study was to investigate the exercise partner and prevalence of constipation. To accumulate further evidence, we aimed to evaluate the association between self-reported physical activity and constipation in a Japanese young population.

## Materials and Methods

### Study population

In this study, we enrolled 12,523 students who had health examination at Ehime University (Ehime, Japan) between April 2015 and April 2017. A specific questionnaire pertaining to constipation was sent to all subjects at the time of their health checkup. After 26 subjects were excluded due to incomplete data, the final analysis sample in this study consisted of 12,497 subjects, all of whom assessed for constipation and exercise habits.

## Ethics statement

All subjects were provided an opt-out, and the study protocol was developed in accordance with the ethical guidelines of the Declaration of Helsinki. This study was approved by the ethics committee of The Ehime University Graduate School of Medicine (approval no. 1610012). All study participants, provided informed written consent prior to study enrollment This study did not include any minors.

### Measurements

Information on smoking, drinking, and medical history was collected using a self-administered questionnaire. Current smoking was defined as positive if a study subject reported smoking. Current drinking was defined as positive if a study subject reported a habit of consuming alcoholic beverages. Height was measured to the nearest millimetre using a stadiometer with the subject standing completely erect. Weight was measured with the subject wearing light clothing. Body mass index (BMI) was calculated as weight in kilograms divided by height in meters squared.

### Assessment of exercise Habits, including presence/absence of exercise partners

We used a self-administered questionnaire to estimate dietary intake using the following questions: (1) Frequency of exercise: “How frequently do you exercise?” (None, 1–2 times per month, 1–3 times per week, and 4 or more times per week), (2) Intensity of exercise: “What is your main type of exercise?” (Light exercise [normal walking, housework, vehicles, ball playing, stretching, etc.], moderate exercise [fast walking, stairlifts, bicycle, softball, hiking, golf, radio calisthenics, etc.], and intense exercise [jogging, climbing stairs, badminton, tennis, mountain climbing, swimming, muscle training, judo, etc.]), and (3) Exercise partner: “Who do you mostly exercise with?” (group, friends, and alone).

### Assessment of constipation

We used a self-administered questionnaire to estimate functional constipation using the following questions: Functional constipation was defined as present if the student answered “yes” to the question “Have you been constipated often recently?”

### Statistical analysis

(1) The frequency of exercise was divided into four categories: none (reference), low: 1–2 times per month, moderate: 1–3 times per week, and high: 4 or more times per week. (2) The intensity of exercise was divided four categories: none (reference), light exercise, moderate exercise, and high intense exercise. (3) The four categories of exercise partners were as follows: none (reference), with a group, with friends, and alone. Estimations of crude odds ratios (ORs) and their 95% confidence intervals (CIs) for constipation in relation to exercise frequency, exercise intensity, and exercise partners were performed using a logistic regression analysis. Multiple logistic regression analyses were used to adjust for potential confounding factors. Age, sex, BMI, drinking, smoking, anemia, and sport injury were selected as potential confounding factors. Statistical analyses were mainly performed using SAS software package version 9.4 (SAS Institute Inc., Cary, NC, USA). All probability values for statistical tests were two-tailed, and p<0.05 was considered statistically significant.

## Results

### Subject Characteristics

Table 1 shows the characteristics of the 12497 study participants. The percentage of men was 60.4% in this cohort. The mean age and BMI were 20.1 years, and 21.37, respectively. The frequency of smoking and drinking were 5.4% and 10.0%, respectively. The prevalence of constipation was 6.5% in this cohort.

**Table 1.**
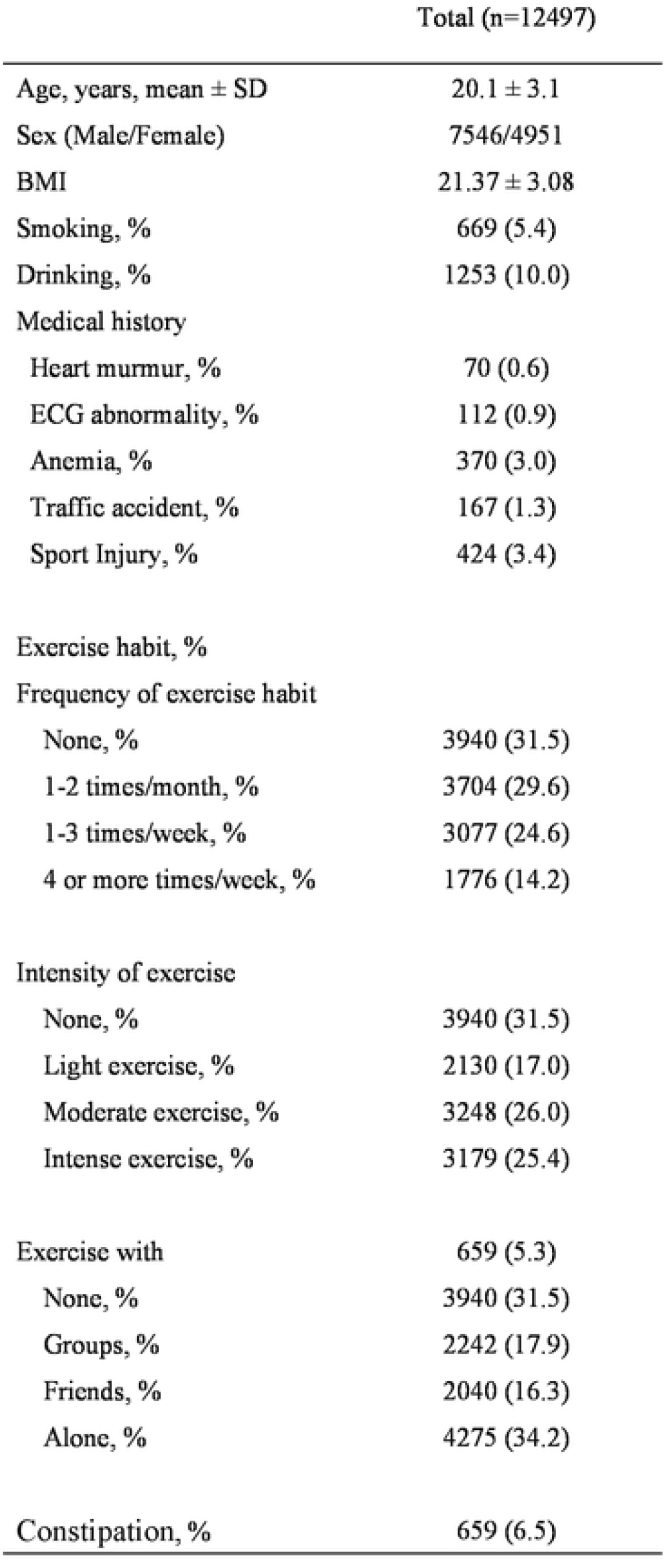
Clinical characteristics of 12497 study participants.

|  | Total (n=12497) |
| --- | --- |
| Age, years, mean ± SD | 20.1 ± 3.1 |
| Sex (Male/Female) | 7546/4951 |
| BMI | 21.37 ± 3.08 |
| Smoking, % | 669 (5.4) |
| Drinking, % | 1253 (10.0) |
| Medical history |  |
| Heart murmur, % | 70 (0.6) |
| ECG abnormality, % | 112 (0.9) |
| Anemia, % | 370 (3.0) |
| Traffic accident, % | 167 (1.3) |
| Sport Injury, % | 424 (3.4) |
| Exercise habit, % |  |
| Frequency of exercise habit |  |
| None, % | 3940 (31.5) |
| 1-2 times/month, % | 3704 (29.6) |
| 1-3 times/week, % | 3077 (24.6) |
| 4 or more times/week, % | 1776 (14.2) |
| Intensity of exercise |  |
| None, % | 3940 (31.5) |
| Light exercise, % | 2130 (17.0) |
| Moderate exercise, % | 3248 (26.0) |
| Intense exercise, % | 3179 (25.4) |
| Exercise with | 659 (5.3) |
| None, % | 3940 (31.5) |
| Groups, % | 2242 (17.9) |
| Friends, % | 2040 (16.3) |
| Alone, % | 4275 (34.2) |
| Constipation, % | 659 (6.5) |

### Association Between Exercise Habits and constipation

Table 2 shows the crude and adjusted ORs and 95% CIs for constipation in relation to exercise habits. The prevalence of constipation among subjects with none, low, moderate, and high frequency of exercise were 7.3%, 4.5%, 4.2%, and 4.2%, respectively. After adjustment for age, sex, BMI, drinking, smoking, anemia, and sport injury, low, moderate, and high frequency of exercise was independently inversely associated with constipation, respectively (adjusted ORs were low: 0.77 [95% CI 0.63–0.94], moderate: 0.75 [95% CI 0.60–0.94] and high: OR 0.70 [95% CI 0.53–0.91], p for trend p=0.002). The prevalence of constipation among low, moderate, and high intensity of exercise was 5.4%, 4. 7%, and 3.2%, respectively. Low, moderate and high intensity of exercise was associated with constipation after adjustment for confounding factors, respectively (adjusted ORs were low: 0.77 [95% CI 0.62–0.97], moderate: 0.77 [95% CI 0.63–0.95] and high: OR 0.68 [95% CI 0.53–0.87], p for trend p=0.001). The prevalence of constipation among exercise with groups, with friends, and alone was 3.8%, 3.0%, and 5.3%, respectively. Exercising with groups and friends was independently inversely associated with constipation (adjusted OR with groups: 0.70 [95% CI 0.53–0.90] and with friends: OR 0.56 [95% CI 0.42–0.74]). The association between exercise alone and constipation was not significant.

**Table 2.** Crude and adjusted odds ratios and 95% confidence intervals for constipation in relation to exercise habits.

| Variable | Prevalence (%) | Crude OR (95% CI) | Adjusted OR (95% CI) |
| --- | --- | --- | --- |
| Constipation |  |  |  |
| Frequency of exercise habit |  |  |  |
| None, % | 289/3940 (7.3) | 1.00 | 1.00 |
| 1-2 times/month, % | 167/3704 (4.5) | 0.60 (0.49–0.73) | 0.77 (0.63–0.94) |
| 1-3 times/week, % | 128/3077 (4.2) | 0.55 (0.44–0.68) | 0.75 (0.60–0.94) |
| 4 or more times/week, % | 75/1776 (4.2) | 0.56 (0.43–0.72) | 0.70 (0.53–0.91) |
| *p* for trend |  |  | 0.002 |
| Intensity of exercise |  |  |  |
| None, % | 289/3940 (7.3) | 1.00 | 1.00 |
| Light exercise, % | 115/2130 (5.4) | 0.72 (0.58–0.90) | 0.77 (0.62–0.97) |
| Moderate exercise, % | 153/3248 (4.7) | 0.63 (0.51–0.76) | 0.77 (0.63–0.95) |
| Intense exercise, % | 102/3179 (3.2) | 0.42 (0.33–0.53) | 0.68 (0.53–0.87) |
| *p* for trend |  |  | 0.001 |
| Exercise with |  |  |  |
| None, % | 289/3940(7.3) | 1.00 | 1.00 |
| Groups, % | 84/2242 (3.8) | 0.49 (0.38–0.63) | 0.70 (0.53–0.90) |
| Friends, % | 61/2040 (3.0) | 0.39 (0.29–0.51) | 0.56 (0.42–0.74) |
| Alone, % | 225/4275 (5.3) | 0.70 (0.59–0.84) | 0.84 (0.70–1.02) |

Odds ratios were adjusted for age, sex, body mass index, drinking, smoking, anemia, and sport injury. OR, odds ratio; CI, confidence interval

**Table 3.** Crude and adjusted odds ratios and 95% confidence intervals for constipation in relation to exercise partner.

| Variable | Prevalence (%) | Crude OR (95% CI) | Adjusted OR (95% CI)* | Adjusted OR (95% CI)** |
| --- | --- | --- | --- | --- |
| Constipation |  |  |  |  |
| Exercise with |  |  |  |  |
| None, % | 289/3940(7.3) | 1.00 | 1.00 | 1.00 |
| Groups, % | 84/2242 (3.8) | 0.49 (0.38–0.63) | 0.70 (0.53–0.90) | 0.78 (0.46–1.33) |
| Friends, % | 61/2040 (3.0) | 0.39 (0.29–0.51) | 0.56 (0.42–0.74) | 0.61 (0.39–0.96) |
| Alone, % | 225/4275 (5.3) | 0.70 (0.59–0.84) | 0.84 (0.70–1.02) | 0.95 (0.64–1.38) |

OR, odds ratio; CI, confidence interval
\*Model 1: Odds ratios were adjusted for age, sex, body mass index, drinking, smoking, anemia, and sport injury.
\*\*Model 2: Odds ratios were adjusted for age, sex, body mass index, drinking, smoking, anemia, sport injury, exercise frequency, and exercise intensity.

## Discussion

In the present study, frequency and intensity of exercise and exercise partner were independently inversely associated with constipation in young Japanese population. To our knowledge, this is the first study to show an inverse association between exercise habit and constipation in the young Japanese population.

Several studies regarding the association between physical activity and constipation have been reported. However, the association between physical activity and constipation was still inconsistent.

In a Swedish study of 15010 subjects aged 45-75 years, the leisure physical activity was significantly inversely associated with constipation [4]. In a Icelandic case control study of 190 children aged 10 year to 18 years, subjects with high frequency of exercise (more than three times a week and/or for more than an hour) was lower onset of functional constipation compared to control patients [5]. In a Chinese study of 1,950 women aged over 50 years, frequency of physical exercise was significantly inversely associated with chronic constipation [6]. In an another Chinese study of 1,698 women who were 37–41 weeks pregnant aged over 35 years, regular exercise was significantly inversely associated with functional constipation [7]. In a Hong Kong study of 32,371 subjects means aged 14.8 years, insufficient exercise was significantly associated with constipation [8]. In a Japanese cohort study of 10,658 subjects, higher physical activity was significantly inversely associated with functional constipation [9]. In a another Japanese study of 5540 adolescents aged 12 to 13 years, physically inactive was significantly associated with constipation [10]. In the third Japanese study, 9,234 subjects aged over 18 years, poor physical activity was significantly associated with constipation [11].

On the other hand, physical activity was not associated with constipation in a US study of 1,069 participants aged 24-77 years [12]. In a British study of 94 primiparous pregnant women aged 19-40 years, no statistically significant differences were identified between light, moderate and vigorous physical activity levels when groups were compared between the constipated and non-constipated [13]. In a Spanish study of 414 participants aged over 50 years, physical activity was not associated with constipation [14]. In a another Spanish study of 415 participants aged 43.8 years, exercise habits were not related to constipation [15]. The discrepancies among these study results may be explained, at least in part, by the differences in the definition of constipation, exercise intensity, race, sample size, study design, assessment of exercise, and age.

The underlying mechanism linking exercise and constipation remains unclear. In particular, the mechanism underlying the beneficial effect of exercising with others remains poorly understood. Additionally, physical activity may enhance intestinal gas clearance [21]. Exercise habits might improve the prevalence of constipation by preventing acid refux, enhancing intestinal gas clearance, and suppressing inflammatory cytokines. Physical activity has been shown to have variable effects on gastrointestinal transit. Physical activity has been shown to have variable effects on gastrointestinal transit. Physical activity promotes bowel movements by improving frequency of bowel activity in the large intestine [22, 23]. In addition, physical activity may change the level of endogenous sex hormones [24], which have been shown to regulate colonic transit time [25]. Physical activity may stimulate appetite, and sufficient dietary intake leads to higher frequency of bowel movement [26]. Exercising with friends and groups might improve functional constipation by reducing depressive symptoms. Further study on the mechanism, especially the association between exercising with others and functional constipation, is needed in the future.

Several limitations of the present study warrant mention. First, the cross-sectional nature of the study does not permit the assessment of causality owing to the uncertain temporality of the association. Second, data on functional constipation in our study was self-reported. We could not estimate gastrointestinal transit time or rectal anal pressure. Therefore, it was not possible to assess the severity of constipation. Similarly, data on participants’ exercise habits was self-reported. The assessment of participants’ exercise habits based on self-administered questionnaire in the present study might not be able to assess the actual physical activity., which might have caused some misclassification bias. However, non-differential misclassification would cause a bias of the odds ratio towards the null. Finally, the patients in the current study might not be representative of the young Japanese population. Given that the present cohort consisted only of university students, it is possible that the relatively high educational status of our population affected health behaviors. In a previous internet study of subjects 40 year older using similar simple questionnaire on constipation, the prevalence of constipation was 51.5% [27].

In conclusion, in the young Japanese population, the frequency and intensity of exercise were independently inversely associated with functional constipation. The inverse association between exercise habits and functional constipation was found in subjects who exercise with friends and in groups but not alone.

## Data Availability

All relevant data are within the manuscript and its Supporting Information files.

## Author Contributions

**Conceptualization:** Junichi Watanabe, Shinya Furukawa

**Data curation:** Junichi Watanabe, Shinya Furukawa

**Formal analysis:** Junichi Watanabe, Shinya Furukawa

**Investigation:** Aki Kato, Katsunori Kusumoto, Yuka Saeki

**Methodology:** Junichi Watanabe, Shinya Furukawa

**Project administration:** Shinya Furukawa, Yoichi Hiasa

**Resources:** Shinya Furukawa, Yoichi Hiasa

**Software:** Junichi Watanabe, Shinya Furukawa

**Supervision:** Yoichi Hiasa

**Validation:** Yasunori Yamamoto, Eiji Takeshita, Yoshio Ikeda, Teruki Miyake, Osamu Yoshida, Naofumi Yamamoto

**Visualization:** Yasunori Yamamoto, Eiji Takeshita, Yoshio Ikeda, Teruki Miyake, Osamu Yoshida, Aki Kato, Katsunori Kusumoto, Yuka Saeki, Naofumi Yamamoto

**Writing – original draft:** Junichi Watanabe, Shinya Furukawa

**Writing – review & editing:** Junichi Watanabe, Shinya Furukawa, Yoichi Hiasa

## Acknowledgments

The authors would like to acknowledge Mikage Oiwa, Hiromi Miyauchi, Yuko Matsumoto, Takako Yamamoto, Hiroko Suzuki, Miyuki Kataoka, Masumi Hino, Tomo Kogama and all of the Health Services Center staff for their support.

